# Estimating timing of pre-emptive livestock vaccination to prevent human Rift Valley fever spillover in Rwanda: a modelling study

**DOI:** 10.64898/2026.11.22.26368714

**Authors:** Edson Rwagasore, Frederic Ntirenganya, Leandre Ishema, Fabrice Ndayisenga, Ziad El-Khatib, Jakob Zinsstag

## Abstract

**Background:** Rift Valley fever recurred in Rwanda in 2018, 2022, 2025 and 2026 despite yearly livestock vaccination. Vaccines protect best if given before transmission speeds up, but the deadline for vaccination and the sectors to prioritise have not been estimated. We aimed to estimate the latest dates to start and finish a campaign and to identify priority sectors.

**Methodology:** We analysed surveillance data from four outbreaks: 1,090 animals and 416 laboratory-confirmed people in 2026, plus 2022, 2025 and 2018 data, vegetation and rainfall records in retrospective observational study. A negative binomial regression of weekly livestock cases gave the growth rate, the basic reproduction number and the start of transmission. We simulated the latest campaign dates 100,000 times, allowing for delays to immunity and campaigns. We ranked 416 sectors using pre-2026 information, tested by leaving out each district in turn. Livestock cases doubled every 9.3 days (95% confidence interval 8.1-10.8) and the basic reproduction number was 1.78 (1.64-1.92); assuming 90% vaccine efficacy, 48.5% coverage (43.4-53.3) was needed to stop spread. For the earliest outbreak (March 2022), live-attenuated vaccine had to start by 11 January (24 December-28 January) and finish by 22 February (11 February-3 March). A campaign begun at first detection would have protected animals after 220 of 1,090 livestock cases (20%) and 149 of 416 human cases (36%) had occurred. Sectors ranked before the season, mainly by the district’s 2022 attack rate, that held 25% of animals contained 66% of livestock cases and 86% of human cases (area under the receiver operating characteristic curve 0.73).

**Conclusions:** Waiting until an outbreak is detected is too late to prevent many early cases, so campaigns should be planned around the earliest-starting outbreak, with a later round to protect animals through the year. Districts and sectors with earlier outbreaks should be prioritised.

**Author summary:** Rift Valley fever is a viral disease of livestock that can spread to people (a zoonosis) through mosquito bites or contact with infected animals. It returned to Rwanda in 2018, 2022, 2025 and 2026 despite yearly livestock vaccination. Vaccines should help most if given before spread begins, but to our knowledge no study had used outbreak data to estimate when and where to vaccinate.

We used animal and human cases from four outbreaks, plus satellite data on plant greenness and rainfall. In 2026, sick animals doubled about every nine days, and spread began in mid-April, before the first sick animal was found. For the earliest outbreak (March 2022), a one-dose vaccine had to be started by 11 January and finished by 22 February. A campaign begun at detection would have taken effect after about one in five animal cases and one in three human cases had already occurred. Areas with earlier outbreaks held most cases, while plant greenness did not help to predict them.

We recommend planning vaccination for January and February each year, with a second round later, starting in those areas. Our dates rest on assumptions, so they should be revisited as more outbreaks are recorded.

## INTRODUCTION

Rift Valley fever (RVF) is a mosquito-borne zoonotic disease that mainly affects cattle, goats and sheep. In animals it causes abortions and newborn deaths. In people it ranges from mild fever to bleeding, brain disease and death [1,2]. Rwanda has had RVF since 2012, with large livestock outbreaks in 2018 and 2022 and further outbreaks in 2024, 2025 and 2026 [3–6]. Genome studies show that the virus stays in Rwanda between outbreaks and is not brought in again each time [6–8]. Outbreaks are also becoming more frequent and spreading to new areas of East Africa [9,10].

Vaccinating animals is the only large-scale way to protect people, because no human vaccine is licensed [11]. But timing matters. One dose of live-attenuated vaccine needs two to three weeks to protect, a two-dose inactivated vaccine needs longer, and a district campaign takes one to two months [12,13]. Vaccinating after the first cases appear is therefore too late to stop the early rise. Models have compared reactive and periodic vaccination and where to send vaccine [14–16], and rainfall, vegetation indices and other risk factors can warn of risk months ahead [17–19]. To our knowledge, however, no study has used outbreak data to set a calendar deadline with stated uncertainty. This study therefore aimed to use surveillance data from four Rwandan outbreaks to estimate when livestock vaccination must start and finish, what earlier vaccination would have prevented, whether this is feasible, and which sectors to prioritise, and to assess whether rainfall or the normalised difference vegetation index (NDVI) can signal when to start.

## METHODS

### Data

We used six line-listed or summary data sets from the Rwanda One Health task force. For 2026 we had 1,090 investigated animals (24 April - 26 August in retrospective observational study accessed from database on 15 September 2026; Rwanda Agriculture and Animal Resources Development Board) and 416 laboratory-confirmed human cases (reverse transcription PCR (RT-PCR) or IgM enzyme-linked immunosorbent assay (ELISA); Rwanda Biomedical Center). For 2022 we had district totals of livestock cases and populations (1,339 cases) and 76 human cases with onset dates. For 2025 we had 161 confirmed animals in Kamonyi and nine human cases. The first-detection date of the 2018 outbreak (May) came from the literature [4,6]. We did not use names, telephone numbers or village names. Human cases were assigned to their district of residence. We also used monthly NDVI (2017-2026, 379 sectors, averaged by district) and monthly district rainfall (2025-2026). The 14 districts with human cases in 2026 were Bugesera, Burera, Gasabo, Gatsibo, Gicumbi, Huye, Kayonza, Kicukiro, Kirehe, Muhanga, Ngoma, Nyarugenge, Ruhango and Rwamagana. This study is a secondary analysis of anonymised routine surveillance data collected under the legal mandates of the Rwanda Biomedical Center (human cases) and the Rwanda Agriculture and Animal Resources Development Board (livestock) during the 2026 Rift Valley fever outbreak response. No participants were recruited, and no names, telephone numbers or village names were used. Formal review board approval was not required. Individual consent was not sought.

### Dates and descriptive analysis

In the 2026 human file, day and month were swapped in every date whose day was 12 or less. For each person we listed both readings of each date, dropped those outside 20 April-31 August 2026, and kept the combination in which onset, consultation, sample and laboratory dates were in order and closest together. This put 98.1% of people in correct order. For human timing we used the laboratory confirmation date, which is complete. Onset date was recorded for 217 people (one onset more than 21 days before confirmation was set to missing) and was not filled in; we compared the share with an onset date across confirmation weeks to check for bias. We coded occupation and exposure from free text (English and Kinyarwanda) with a keyword list checked on a random sample. ‘Contact with animals or animal products’ combines the direct-contact, meat and milk groups. We report counts, percentages and median with interquartile range. We compared Ngoma with all other districts using the ratio of human to livestock cases (odds ratio with Woolf 95% confidence interval and Fisher’s exact test); we did the same for 2022 using district totals. This is descriptive because case-finding effort was not recorded. A livestock record was counted as vaccinated if marked ‘Yes’.

### Growth and reproduction numbers

We counted livestock cases by week (weeks start on Monday; ISO week 17 starts on 20 April) and fitted a negative binomial regression, log E[Yt] = α + βt, by maximum likelihood from the first week to the peak week (ISO weeks 17-28, 12 weeks including one week with no cases) [20,21]. The daily growth rate is r = β/7 and the doubling time is ln 2/r. The 2022 human onset curve was fitted the same way, and its peak week was taken from the onset dates. The basic reproduction number was R0 = (1 + rθ) k, where the generation interval is gamma distributed with mean μ = 8 days and standard deviation 3 days (k = (μ/σ)2, θ = σ2/μ).24 Herd immunity is 1 - 1/R0, and the minimum coverage is this value divided by vaccine efficacy (0.90). We repeated the calculation with mean generation intervals of 16 and 20 days (standard deviation 0.375 × mean). We estimated the effective reproduction number Rt from daily livestock counts with a 14-day sliding window, a gamma serial interval (mean 8, standard deviation 3 days) and a Gamma (1, 5) prior, and reported windows with at least 20 cases [22–24].

### Start of transmission and campaign deadlines

We traced the fitted curve back to the date at which one case per week is expected (a simple extrapolation of the fitted line, not the non-parametric back-projection of Becker and colleagues [25]). Transmission start T0 is this date minus the delay from infection to detection: incubation (uniform 2-6 days) plus time to investigation (uniform 0-5 days). We also assumed that only 25% of early cases were found, which moves T0 earlier by ln(1/0.25)/r days, and repeated the analysis with an investigation delay of 0-14 days, with incubation of 2-10 days, and with no delay. The latest completion date is T0 minus the time to immunity, which we took as uniform 14-21 days for live-attenuated vaccine and 28-49 days for two doses 4 weeks apart. The latest start date is the completion date minus campaign time (uniform 28-56 days). We drew 100,000 values of (α, β) from their joint normal distribution and of every delay, as in earlier timing simulations [26,27], and report medians with 2.5^th^-97.5th percentiles.

### Other outbreaks

For 2018 and 2022 we took the first-detection date from reports (15-31 May 2018; 21 March 2022) [3] and subtracted the time from transmission start to first detection estimated for 2026 (median 9 days). For 2025, human illness was recognised before animals were tested, so we set T0 to the first human onset (23 November) minus 21-50 days, the gap between transmission start and first human onset in 2022 (21 days) and 2026 (50 days). Deadlines were then calculated as above.

### Earlier vaccination and feasibility

With vaccine coverage c (animals vaccinated and protected in time) and vaccine efficacy 0.9, the reproduction number is R0(1 - 0.9c). When it is below 1, the expected size of an outbreak started by I0 = 3 cases (the first-week count) is I0/(1 - Reff); we multiplied livestock cases by the observed human-to-livestock ratio (416/1,090) to estimate human cases. For a campaign begun at first detection (24 April), we added the same immunity and delivery times and counted the share of cases that came later (livestock by investigation date, people by laboratory confirmation date). For feasibility we used 2022 district livestock populations and a reported vaccination rate of 112,110 animals in 51 days in Ngoma [5], and divided the animals to be vaccinated by this rate for one team or for seven district teams in parallel.

### Rainfall and vegetation

NDVI anomalies are z-scores against the 2017-2025 mean and standard deviation of each calendar month, averaged over the 14 affected districts; we also averaged them over January-April of each year. An alert was a monthly anomaly above +1. We looked at the five months before each first detection, and counted an alert month in 2017-2025 as a false alarm if no first detection followed within five months. Rainfall was available for 2025 and 2026 only, so we compared January-April of the two years.

### Sectors and vegetation zones

We used the administrative sector as the unit of place: the sector of the animal for livestock cases and the sector of residence for people. We analysed the 416 sectors with NDVI records. Animals per sector are not recorded, so we spread each district’s ruminant population (cattle, goats and sheep in the 2022 outbreak file) equally over its sectors. For each sector we calculated the mean NDVI and the monthly NDVI profile for 2017-2025, and the NDVI anomaly for January-March 2026. We grouped sectors into three vegetation zones by k-means clustering of the 12-month profile, and compared livestock cases per 1,000 animals and human cases per 100,000 animals between zones. We estimated the rate ratio for livestock cases per standard deviation of mean NDVI with a quasi-Poisson regression with log animals as offset, with and without district as a fixed effect.

### Predicting hot spots and targeting

To rank sectors before the season, we fitted quasi-Poisson regressions of 2026 livestock cases per sector, with log animals as offset, on the 2022 district attack rate (log of one plus cases per 1,000 ruminants), a 2022 human case in the sector and a 2025 animal or human case in the sector. A third model added long-term NDVI and the January-March 2026 anomaly. We tested each model by leaving out one district at a time, fitting the others and predicting the held-out district. We report the area under the receiver operating characteristic curve for finding sectors with livestock cases, and the share of 2026 livestock and human cases in the top-ranked sectors that hold 10% or 25% of animals; the best possible ranking orders sectors by 2026 cases per animal. To estimate the effort needed, we ranked sectors by 2026 cases per animal, found the top-ranked sectors that hold 50% and 80% of livestock cases and the share of animals in them, and set coverage inside them at 80%, so that total coverage is 0.8 times that share. We compared this with blanket coverage c, which gives a reproduction number of R0(1 - 0.9c). We also listed the 15 sectors with the most livestock cases. The ranked list of all sectors is provided with the code.

### Software

All analyses used R 4.5.1 version (packages readxl, ggplot2 and patchwork) with a fixed random seed. This study followed the Strengthening the Reporting of Observational Studies in Epidemiology (STROBE) guideline; the completed checklist is provided as S1 Checklist.

## RESULTS

From 24 April to 26 August 2026, we investigated 1,090 animals in 20 districts; 956 were cattle and 12.7% died (Table 1). Between 10 June and 10 August, 416 people in 14 districts were confirmed by laboratory test: 377 (90.6%) lived in the Eastern Province and 270 (64.9%) in Ngoma, and 10.1% died. Most were farmers or herders (77.8%), and 70.7% reported contact with animals or eating or drinking animal products. Ngoma had 71.4 human cases per 100 livestock cases, against 20.5 elsewhere (odds ratio 3.48, 95% CI 2.75-4.41) and 5.5 in 2022 (Table 1). Livestock cases peaked in the week of 6 July (n = 264). Laboratory confirmation of people began on 10 June, 47 days after the first animal case, and peaked in the week of 15 June (n = 109; Fig. 1a). An onset date was recorded for 52% of people but for only 12% of those confirmed in the week of 15 June, so we used the laboratory date for human timing (Table 1).

**Figure 1.**
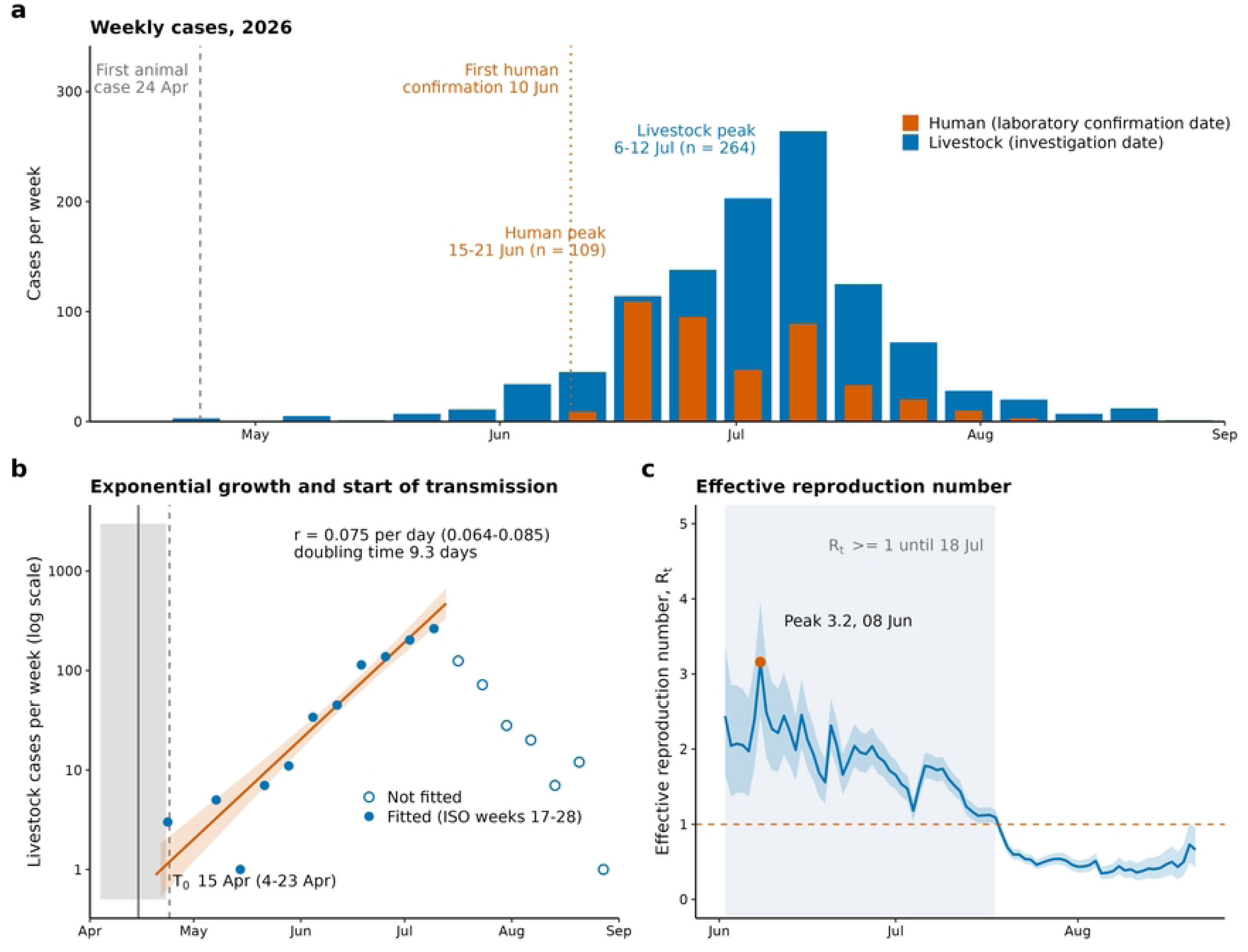
The 2026 outbreak. **a**, Weekly livestock cases (blue, investigation date) and human cases (orange, laboratory confirmation date); the dashed line is the first animal case and the dotted line the first human confirmation. **b,** Livestock cases per week on a log scale with the negative binomial fit to ISO weeks 17-28 (filled points; band, 95% CI) and the estimated start of transmission T0 (grey; line, median; box, 95% interval); the dashed line is the first animal case. **c,** Effective reproduction number Rt (14-day window; blue band, 95% credible interval; dashed line, Rt = 1); shading marks the period with Rt at or above 1.

**Table 1.** Characteristics and sector distribution of livestock and human cases, Rwanda, 2026.

| **Characteristic** | **Livestock (n = 1090)** | **Human (n = 416)** |
|---|---|---|
| Species: cattle / goat / sheep | 956 / 125 / 9 | – |
| Female | 848 (77.8%) | 134 (32.2%) |
| Age, median (IQR), years | – | 36 (24-47) |
| Districts affected (of 30) | 20 | 14 |
| Eastern Province | 768 (70.5%) | 377 (90.6%) |
| Northern Province | 128 (11.7%) | 16 (3.8%) |
| Southern Province | 105 (9.6%) | 9 (2.2%) |
| Kigali City | 89 (8.2%) | 14 (3.4%) |
| Ngoma District | 378 (34.7%) | 270 (64.9%) |
| Kirehe District | 128 (11.7%) | 56 (13.5%) |
| Kayonza District | 168 (15.4%) | 35 (8.4%) |
| Died (case fatality) | 138 (12.7%) | 42 (10.1%) |
| Occupation (n = 387 recorded): farmer or herder | – | 301 (77.8%) |
| veterinary or animal health | – | 20 (5.2%) |
| child or student | – | 27 (7.0%) |
| butcher or other | – | 39 (10.1%) |
| Exposure: direct animal contact | – | 168 (40.4%) |
| meat consumption or handling | – | 96 (23.1%) |
| milk consumption | – | 52 (12.5%) |
| unknown or none identified | – | 85 (20.4%) |
| **Sectors (416 analysed)** |  |  |
| Sectors with at least one case | 151 | 74 |
| Cases in the 21 sectors with the highest livestock case rates (5.0% of sectors; 3.7% of animals) | 530 (50.2%) | 185 (44.9%) |
| Cases in the 63 sectors with the highest livestock case rates (15.1% of sectors; 12.5% of animals) | 845 (80.1%) | 350 (85.0%) |
| Cases in the 15 sectors with the most livestock cases (3.6% of sectors; 3.7% of animals) | 509 (48.2%) | 251 (60.9%) |
| Cases in the 54 sectors with a case in 2022 or 2025 (13.0% of sectors; 14.0% of animals) | 330 (31.3%) | 183 (44.4%) |

*Values are n (%) unless stated. Human cases are by district of residence. Occupation percentages use people with a recorded occupation; exposure groups can overlap. IQR, interquartile range. Sector rows use the 416 sectors with vegetation records, which held 1,055 of 1,090 livestock cases and 412 of 416 human cases; percentages are of the cases in these sectors, human cases are by sector of residence, and animal shares assume animals are equal across the sectors of a district. Human cases per 100 livestock cases were 71.4 in Ngoma and 20.5 elsewhere (odds ratio 3.48, 95% CI 2.75-4.41), and 5.5 in Ngoma in 2022. An onset date was recorded for 52% of people and for 12% of those confirmed in the week of 15 June.*

### Growth and transmissibility

Weekly livestock cases rose exponentially to the peak (r = 0.075 per day, 95% CI 0.064-0.085), doubling every 9.3 days (8.1-10.8; Fig. 1b). With a mean generation interval of 8 days, the basic reproduction number R0 was 1.78 (1.64-1.92). If the vaccine is 90% effective, 48.5% (43.4-53.3%) of animals must be vaccinated to stop spread (Table 2). Mosquitoes may lengthen the generation interval; at 16-20 days R0 was 3.0-3.9 and the coverage needed was 74-83%. We therefore treat 49% as a minimum and aim for at least 80%. In 2022 human cases grew more slowly (R0 1.4, 1.2-1.6) and peaked in the week of 30 May, about six weeks after the animal peak on 14 April 3. The effective reproduction number peaked at 3.2 (2.4-4.0) on 8 June, stayed at or above 1 until 18 July, and then fell below 1 (Fig. 1c).

**Table 2.** Epidemic estimates and latest dates to start and finish a vaccination campaign.

| Outbreak (first detection) | Transmission start | Live-attenuated: start by | Live-attenuated: finish by | Two-dose: start by | Two-dose: finish by |
|---|---|---|---|---|---|
| **2022** (21 Mar) | 12 Mar (1-20 Mar) | **11 Jan** (24 Dec-28 Jan) | **22 Feb** (11 Feb-3 Mar) | **21 Dec** (30 Nov-11 Jan) | **1 Feb** (17 Jan-15 Feb) |
| 2026 (24 Apr) | 15 Apr (4-23 Apr) | 14 Feb (27 Jan-3 Mar) | 28 Mar (17 Mar-6 Apr) | 24 Jan (2 Jan-14 Feb) | 7 Mar (20 Feb-21 Mar) |
| 2026, if only 25% of early cases were found | 27 Mar (14 Mar-6 Apr) | 26 Jan (7 Jan-14 Feb) | 10 Mar (24 Feb-20 Mar) | 5 Jan (13 Dec-28 Jan) | 16 Feb (30 Jan-4 Mar) |
| 2018 (late May) | 13 May (30 Apr-26 May) | 15 Mar (23 Feb-4 Apr) | 26 Apr (12 Apr-9 May) | 22 Feb (30 Jan-17 Mar) | 5 Apr (19 Mar-22 Apr) |
| 2025 (23 Nov) | 18 Oct (4 Oct-1 Nov) | 20 Aug (28 Jul-11 Sep) | 30 Sep (16 Sep-15 Oct) | 30 Jul (4 Jul-24 Aug) | 10 Sep (21 Aug-29 Sep) |
| **Epidemic estimates behind the deadlines and the effort needed** |  |  |  |  |  |
| Growth rate per day; doubling time (days) | 0.075 (0.064-0.085); 9.3 (8.1-10.8) |  |  |  |  |
| Basic reproduction number R0 at generation interval 8 days; at 16 / 20 days | 1.78 (1.64-1.92); 3.02 / 3.89 |  |  |  |  |
| Vaccine coverage needed at 90% efficacy (%), generation interval 8 days; at 16 / 20 days | 48.5 (43.4-53.3); 74.3 / 82.5 |  |  |  |  |
| Peak Rt (date); last day with Rt of 1 or more | 3.2 (2.4-4.0) on 08 Jun 2026; 18 Jul 2026 |  |  |  |  |
| Days from transmission start to first animal case; median start across the four delay assumptions | 9 (1-19); 08 Apr 2026 - 21 Apr 2026 |  |  |  |  |
| R0 from 2022 human onsets | 1.39 (1.21-1.58) |  |  |  |  |
| Other outbreaks | 2022: 1,339 livestock cases (district totals). 2025: 161 animals and 9 people; first human onset on 23 Nov 2025, 24 days before the first animal confirmation |  |  |  |  |
| If 60% / 80% of animals were vaccinated and protected at transmission start: reproduction number; expected livestock cases; expected human cases (observed 1,090 and 416) | 0.82; 17; 6 / 0.50; 6; 2 |  |  |  |  |
| Campaign begun at first detection: animals protected from; cases up to that date | 22 Jun 2026; livestock 220 of 1090 (20%), human 149 of 416 (36%) |  |  |  |  |
| Vaccinating 80% of the Eastern Province cattle (513,338 cattle; 410,670 animals) at 2,198 animals a day: days for one team / seven district teams | 187 / 27 |  |  |  |  |
| **Vegetation and rainfall (14 affected districts)** |  |  |  |  |  |
| NDVI anomaly, mean of January-April 2026 (z-score) | 0.86, the highest of 2017-2026 |  |  |  |  |
| First month of 2026 with NDVI anomaly above +1 | March |  |  |  |  |
| Alert months 2017-2025 (anomaly above +1); followed by a first detection within five months | 15; 4 |  |  |  |  |
| Rainfall, January-April 2025 / 2026 (mm); change | 430 / 509; +18% |  |  |  |  |

*Median (95% interval). Dates are in the year of first detection, except December starts, which fall in the previous year. †First-detection date from the literature only. Transmission start set from the first human onset because people fell ill before animals were tested. The earliest outbreak (2022, bold) sets the planning calendar. In the lower blocks, intervals for growth and reproduction numbers are confidence intervals from the negative binomial fit and intervals for dates come from the simulations; expected cases assume three early cases and the observed ratio of human to livestock cases; the NDVI anomaly is a z-score against 2017-2025.*

### When transmission started

Tracing the growth curve back, and allowing for incubation (2-6 days) and time to investigation (0-5 days), transmission started on about 15 April 2026 (4-23 April), nine days (1-19) before the first animal was recorded (Table 2). The first human onset on record came 50 days later (4 June) and the first laboratory confirmation 56 days later (10 June). If only a quarter of early cases were found, transmission started on 27 March. Other delay assumptions moved the start to between 8 and 21 April (Table 2).

### Campaign deadlines

We subtracted the time to immunity (2-3 weeks for live-attenuated vaccine, 4-7 weeks for two doses) and the time to run a campaign (4-8 weeks). For 2026, live-attenuated vaccination had to start by 14 February (27 January-3 March) and finish by 28 March (Table 2). The 2022 outbreak, first found on 21 March, set an earlier deadline: start by 11 January (24 December-28 January) and finish by 22 February (11 February-3 March); two-dose vaccine had to start by 21 December. The 2022 animal file gave district totals only (1,339 cases), so we used its first-detection date (Table 2). The 2025 outbreak in Kamonyi (161 animals, 9 people) was found through human illness that began on 23 November, 24 days before the first animal was confirmed, and gave the latest deadline (live-attenuated start by 20 August; Fig. 2). The 2018 date comes from the literature alone (start by 15 March).

**Figure 2.**
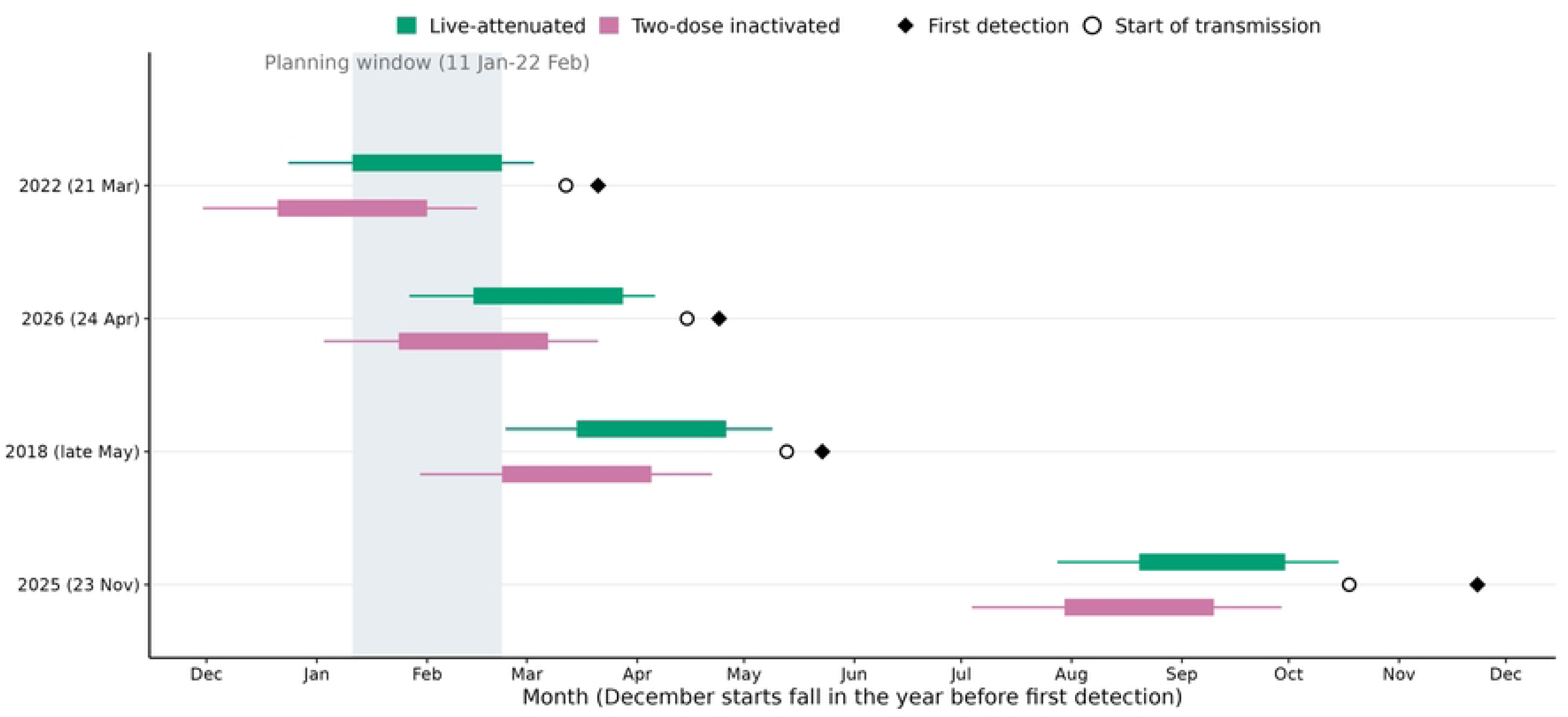
Latest campaign dates by outbreak. Bars run from the median latest start to the median latest finish for live-attenuated (green) and two-dose (pink) vaccine; thin lines show 95% intervals, diamonds the first detection and open circles the estimated start of transmission. The grey band is the planning window set by the earliest outbreak (2022; live-attenuated vaccine, 11 January-22 February).

### What earlier vaccination would have changed

If 60% or 80% of animals had been vaccinated and protected when transmission started, the reproduction number would have fallen to 0.82 or 0.50. A chain begun by three early cases would then be expected to cause about 17 or 6 livestock cases instead of 1,090, and about 6 or 2 human cases instead of 416 (Table 2). These are best-case figures. A campaign begun at first detection would instead have protected animals from about 22 June (live-attenuated), after 20% of livestock cases and 36% of human cases. Vaccinating 80% of the 513,338 cattle in the Eastern Province (410,700 animals) at the rate reported for a recent Ngoma campaign (2,198 animals a day) would take 187 days for one team, but 27 days for seven district teams working together (Table 2).

### Where cases clustered

We analysed the 416 sectors with vegetation records, which held 1,055 of 1,090 livestock cases and 412 of 416 human cases. Livestock cases came from 151 of these sectors and human cases from 74. The 21 sectors (5%) with the highest case rates held half of the livestock cases, and 63 sectors (15%) held 80% of livestock cases and 85% of human cases (Table 1). The 15 sectors with the most livestock cases, mostly in Ngoma and Kayonza, held 48% of livestock cases and 61% of human cases (Table 1; Fig. 4c). Sectors with a case in 2022 or 2025 made up 13% of sectors but held 31% of livestock cases and 44% of human cases (Table 1).

### Rainfall and vegetation

The 2026 season was green and wet. NDVI averaged 0.86 standard deviations above normal from January to April, the highest since 2017, and rainfall was 18% above 2025 (Table 2) (Fig. 3). But NDVI first passed +1 in March 2026, after the 14 February start deadline. An alert at +1 would have fired in 15 months of 2017-2025, and only 4 of them, all before the 2025 outbreak, were followed by a detection within five months. NDVI was not raised in the five months before the 2018 or 2022 outbreaks.

**Figure 3.**
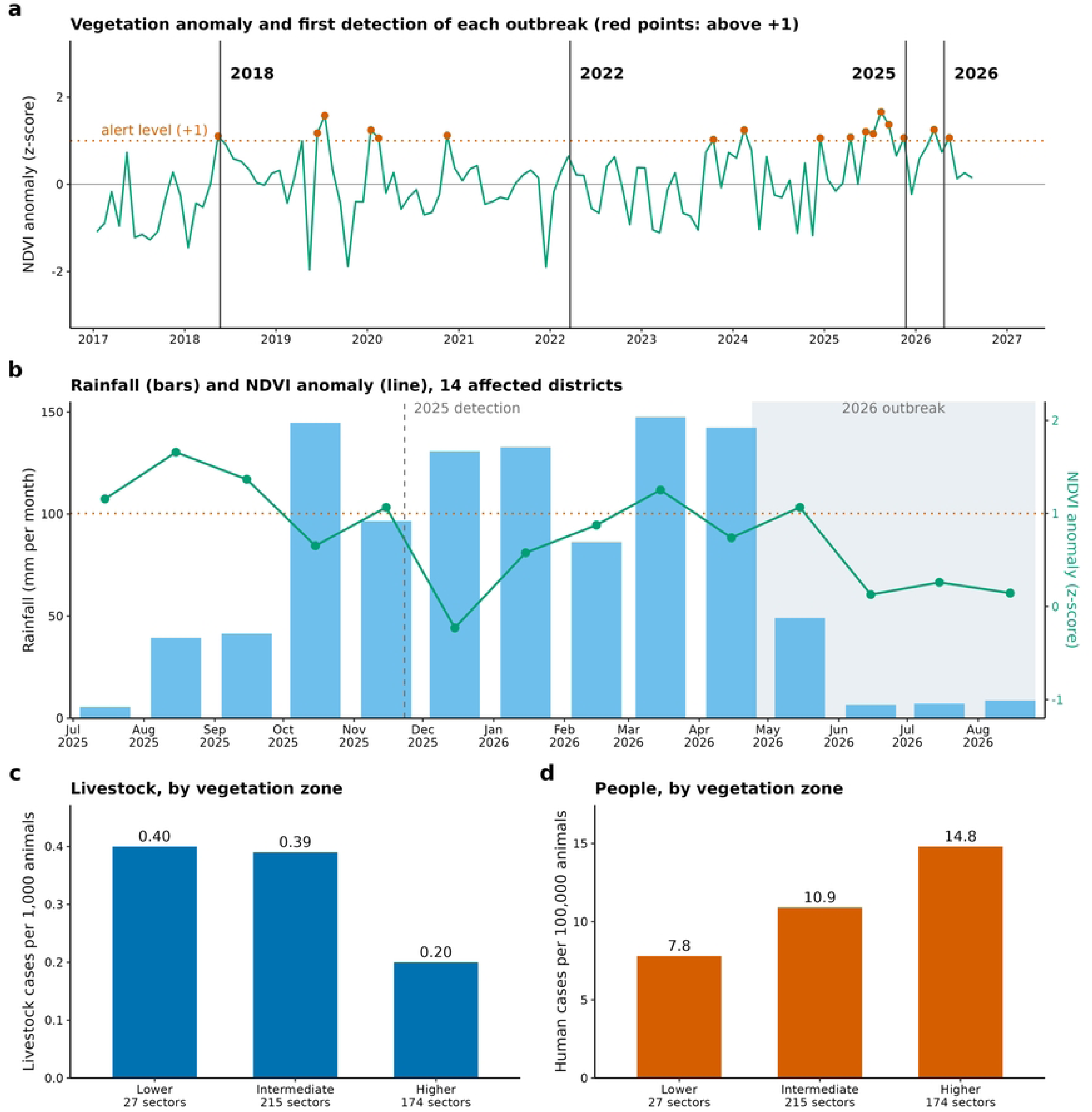
Vegetation, rainfall and vegetation zones. a, NDVI anomaly (z-score against 2017-2025 for each calendar month) averaged over the 14 affected districts, with the first detection of each outbreak; the dotted line is the +1-alert level and red points are months above it. b, Monthly rainfall (blue bars) and NDVI anomaly (green line), July 2025-August 2026; grey shading marks the 2026 outbreak and the dashed line the 2025 detection. c, d, Livestock cases per 1,000 animals and human cases per 100,000 animals in three vegetation zones, with sectors grouped by their 2017-2025 monthly NDVI profile (mean NDVI shown). Rate ratio for livestock cases per standard deviation of mean NDVI: 0.73 (95% CI 0.54-1.00) across all sectors and 1.30 (1.09-1.55) within districts.

We grouped sectors into three vegetation zones by their monthly NDVI profile for 2017-2025 (Fig. 3c,d). Livestock cases per 1,000 animals were 0.40, 0.39 and 0.20 from the lower to the higher NDVI zone, and human cases per 100,000 animals were 7.8, 10.9 and 14.8. Across all sectors, greener sectors had fewer livestock cases (rate ratio per standard deviation of NDVI 0.73, 95% CI 0.54-1.00), but within districts the association reversed (1.30, 1.09-1.55; Fig. 3 legend). NDVI therefore did not separate high-risk from low-risk sectors.

### Predicting hot spots before the season

We ranked sectors with information from before 2026 and tested the ranking by leaving out each district in turn. The ranking identified sectors with livestock cases with an area under the receiver operating characteristic curve (AUC) of 0.73, against 0.50 for random choice (Fig. 4b). Most of the predictive value came from the district’s 2022 attack rate; sector history added little (AUC 0.72 with the district rate alone). The top-ranked sectors that held 10% of animals contained 56% of livestock cases and 82% of human cases, and those that held 25% of animals contained 66% and 86% (Fig. 4a). The best possible ranking, by the 2026 cases themselves, is shown by the dashed lines. Adding vegetation lowered the AUC to 0.70 (Fig. 4b).

**Figure 4.**
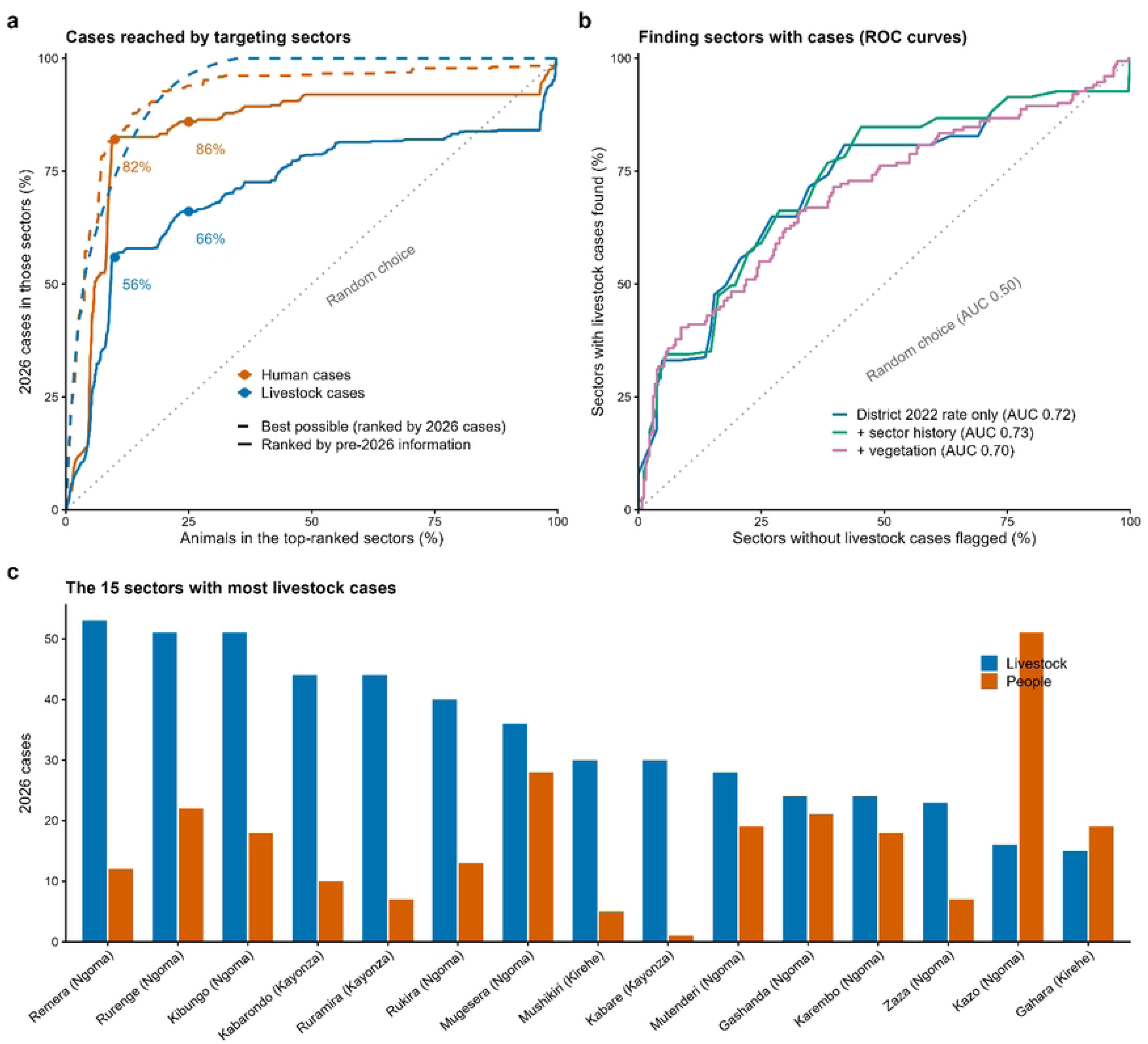
Targeting sectors. a, Share of 2026 cases in the top-ranked sectors against the share of animals in those sectors. Solid lines rank sectors with information from before 2026 (2022 district attack rate and sector history) and are tested by leaving out each district in turn; dashed lines rank sectors by 2026 cases (best possible); the dotted line is random choice; points mark 10% and 25% of animals. b, Receiver operating characteristic curves for finding sectors with livestock cases, with the area under each curve (AUC); this is a different measure from panel a. c, The 15 sectors with the most livestock cases in 2026, with human cases by sector of residence.

### Targeting the most affected sectors

In hindsight, with sectors chosen by their 2026 case rates and animals assumed equally spread within districts, the 21 sectors that held half of livestock cases hold 3.7% of animals and the 63 sectors that held 80% hold 12.5% (Table 1). Vaccinating 80% of animals in those 63 sectors would need a total coverage of 10% (0.8 x 12.5%). This is an upper bound, because the sectors were chosen with 2026 data: the pre-season ranking reached 56% of livestock cases at 10% of animals (Fig. 4a). Blanket coverage of 25% would leave the reproduction number at 1.38 (1.78 x (1 - 0.9 x 0.25)), above 1, and the blanket threshold stays at 48.5% (Table 2).

## DISCUSSION

Vaccinating animals before transmission starts could prevent many early RVF cases, but the window is short. In 2026 transmission started in mid-April, the first person fell ill about seven weeks later and animal cases peaked about twelve weeks later. A campaign begun at first detection would have protected animals only after a fifth of livestock cases, so it shortens the tail but cannot stop the rise. This fits models in which periodic vaccination of cattle, with reactive vaccination added, works best [15].

The deadline depends on the earliest likely start, not on one outbreak. First detections ranged from 21 March to 23 November. Using the earliest, we suggest a planning calendar: start live-attenuated vaccination in early January, finish by mid-to-late February at 80% or more coverage, and start two-dose vaccination by mid-December. This is a planning estimate from four outbreaks, not a fixed date, and should be revised each year. Because of the November 2025 outbreak, protection from a February campaign must last eight to ten months or be topped up by a mid-year round. How long protection lasts in the field is unknown and should be measured by blood surveys. New vaccines that protect several species may change these time lags, and the dates should then be updated [28].

Rainfall and vegetation did not give a usable trigger. The high NDVI of 2026 crossed the alert line too late, and most alerts were false. This agrees with genome evidence that the virus persists in Rwanda [6,7,29], so risk is present every year and not only after floods. NDVI and rainfall could still help decide how hard to push in unusually wet years [17,18].

Cases clustered, and places hit before were hit again: Ngoma and Kirehe had the highest district attack rates in 2022 and were among the three districts with most livestock cases in 2026, which fits local persistence of the virus [6,7,29]. Ranking sectors before the season placed two thirds of livestock cases and most human cases in sectors holding a quarter of animals, so limited doses can go there first. Vegetation did not help. Targeting lowers cases only if transmission stays mostly within sectors; if animals and mosquitoes mix widely, the 48.5% blanket threshold still applies [30,31].

Human laboratory confirmations peaked before animal investigations (week of 15 June against 6 July). This most likely reflects when people were tested: testing began on 10 June, and onset dates are missing for most people confirmed in mid-June, so the true timing of illness cannot be rebuilt. In 2022 people peaked about six weeks after animals, and in 2025 people fell ill before animals were tested. People are therefore found earlier than infected animals, but animals are still the source. The high human-to-animal ratio in Ngoma was not seen in 2022, so it probably reflects how hard cases were looked for. Most patients were farmers or herders, as in other East and Central African studies,21 and no human vaccine exists [11,32]. Strengths are the first-detection dates from four outbreaks, detailed animal and human data from 2026, sensitivity analyses of key assumptions, and a sector ranking tested on districts left out of the fitting.

### The study has limits

Growth came from animals that were investigated, and more effort later in the outbreak would raise growth and R0. The start of transmission depends on assumed delays and on how many early cases were found. We applied the 2026 detection delay to other outbreaks and used one published date for 2018. Generation intervals and campaign times were assumed. The 44% (479 of 1,090) of animals had a vaccination record (85% of these recorded as vaccinated, with no product or date), so we could not judge how well the vaccine worked. Sector rankings rest on one season of case counts, which also reflect where cases were looked for, and we spread each district’s animals equally across its sectors.

## Data Availability

The datasets and script are shared during submission of the manuscript as zip file.

## Acknowledgements

We thank the field epidemiologists, veterinary officers, laboratory scientists and members of the Rwanda One Health task force whose surveillance produced these data.

## References

1. Rostal MK, Thompson PN, Anyamba A, Bett B, Cêtre-sossah C, Chevalier V, et al. Rift Valley fever epidemiology : shifting the paradigm and rethinking research priorities. Lancet Planet Heal 2025. 2025;9: 1–10. doi:10.1016/j.lanplh.2025.101299

2. Rostal MK, Prentice JC, Ross N, Kemp A, Peter N, Anyamba A, et al. Localized Rift Valley fever virus persistence explains epidemic and interepidemic dynamics and guides control strategies. royalsocietypublishing.org/. 2026. Proc R Soc B 2025; 292: 20250453. doi:10.1098/rspb.2025.0453

3. Remera E, Rwagasore E, Nsekuye O, Semakula M, Gashegu M, Rutayisire R, et al. Rift Valley Fever Epizootic, Rwanda, 2022. Emerg Infect Dis. 2024;30: 2022–2024. 10.3201/eid3010.240264

4. Ishema L, Colombe S, Ndayisenga F, Uwibambe E, Van E, Shyaka A, et al. One Health investigation and response to a nationwide outbreak of Rift Valley fever in Rwanda - March to December 2022. One Health. 2024;19:100854. doi:10.1016/j.onehlt.2024.100854.

5. Niyonzima E, Nkurunziza F, Ngaboyimana JD, Nyirandahiriwe V, Mvuyekure F, Ndayisenga F, et al. Investigation and response to Rift Valley fever outbreak in ruminant livestock from Ngoma District, eastern province of Rwanda, 2024. One Health. 2026;22:101332. doi: 10.1016/j.onehlt.2026.101332

6. Nsengimana I, Juma J, Roesel K, Gasana MN, Ndayisenga F, Muvunyi CM, et al. Genomic Epidemiology of Rift Valley Fever Virus Involved in the 2018 and 2022 Outbreaks in Livestock in Rwanda. Viruses. 2024; 1–16;16(7):1148. doi: 10.3390/v16071148.

7. Juma J, Tegally H, Konongoi SL, Roesel K, Sang R, Oliviera T De, et al. Spatiotemporal patterns of Rift Valley fever virus in Africa : a retrospective genomic epidemiology and phylodynamic modelling study. Lancet Microbe. 2026;7(3):101251. doi: 10.1016/j.lanmic.2025.101251.

8. Clark MHA, Warimwe GM, Di Nardo A, Lyons NA, Gubbins S. Systematic literature review of Rift Valley fever virus seroprevalence in livestock, wildlife and humans in Africa from 1968 to 2016. PLoS Negl Trop Dis. 2018;12: 1–19. doi:10.1371/journal.pntd.0006627

9. Umutoni B, Zakham F, Ndayambaje M, Hewins B, Gasana MN, Ostadgavahi AT, et al. Rift Valley Fever outbreaks in the East African Community : insights from ProMed data (. Front Public Heal. 2024. 10.3389/fpubh.2024.1298594

10. Situma S, Nyakarahuka L, Omondi E, Mureithi M, Mweu MM, Muturi M, et al. Widening geographic range of Rift Valley fever disease clusters associated with climate change in East Africa. BMJ Glob Heal. 2024. doi:10.1136/bmjgh-2023-014737

11. Jenkin D, Wright D, Folegatti PM, Platt A, Poulton I, Lawrie A, et al. Safety and immunogenicity of a ChAdOx1 vaccine against Rift Valley Fever in adults : an open-label, non-randomised, first-in-human phase 1 clinical trial. Lancet Infect Dis. 2023;23: 956–964. doi:10.1016/S1473-3099(23)00068-3.Safety

12. Bird BH, Nichol ST. Breaking the chain : Rift Valley fever virus control via livestock Brian H Bird and Stuart T Nichol. Curr Opin Virol. 2012;2: 315–323. doi:10.1016/j.coviro.2012.02.017

13. Alkan C, Jurado-cobena E. Advancements in Rift Valley fever vaccines : a historical overview and prospects for next generation candidates. NPJ Vaccines. 2023; 1–16. doi:10.1038/s41541-023-00769-w

14. Id AD, Ezanno P, Id BD. Mechanistic models of Rift Valley fever virus transmission : A systematic review. 2022. doi:10.1371/journal.pntd.0010339

15. Gachohi JM, Njenga MK, Kitala P, Bett B. Modelling Vaccination Strategies against Rift Valley Fever in Livestock in Kenya. PLoS Negl Trop Dis. 2016; 1–22. doi:10.5061/dryad.779rd

16. Tennant WSD, Cardinale E, Moutroifi Y, Spencer SEF, Charafouddine O, Tildesley MJ, et al. Effectiveness and equity of vaccination strategies against Rift Valley fever in a heterogeneous landscape. PLoS Negl Trop Dis. 2025;2025: 1–24. doi:10.1371/journal.pntd.0013346

17. Anyamba A, Chretien J, Small J, Tucker CJ, Formenty PB, Richardson JH, et al. Prediction of a Rift Valley fever outbreak. Proc Natl Acad Sci. 2009;106: 955–959; 10.1073/pnas.0806490106

18. Linthicum KJ, Anyamba A, Tucker CJ, Kelley PW, Myers MF, Peters CJ. Climate and satellite indicators to forecast Rift Valley fever epidemics in Kenya. Science. 1999;285(5426):397–400; 10.1126/science.285.5426.397

19. Métras R, Jewell C, Porphyre T, Thompson PN, Pfeiffer DU, Collins LM, et al. Risk factors associated with Rift Valley fever epidemics in South Africa in 2008-11. Sci Rep. 2015;5: 1–7. doi:10.1038/srep09492

20. Kucharski AJ, Russell TW, Diamond C, Liu Y, Edmunds J, Funk S, et al. Early dynamics of transmission and control of COVID-19 : a mathematical modelling study. Lancet Infect Dis. 2020; 553–558. doi:10.1016/S1473-3099(20)30144-4

21. Camacho A, Kucharski AJ, Funk S, Breman J, Piot P, Edmunds WJ. Potential for large outbreaks of Ebola virus disease. Epidemics. 2014. doi:10.1016/j.epidem.2014.09.003

22. Cori A, Ferguson NM, Fraser C, Cauchemez S. A New Framework and Software to Estimate Time-Varying Reproduction Numbers During Epidemics. Am J Epidemiol. 2013;178: 1505–1512. doi:10.1093/aje/kwt133.

23. Thompson RN, Stockwin JE, van Gaalen RD, Polonsky JA, Kamvar ZN, Demarsh PA, et al. Improved inference of time-varying reproduction numbers during infectious disease outbreaks. Epidemics. 2019;29:100356. doi: 10.1016/j.epidem.2019.100356.

24. Id RKN, Bhatt S, Cori A, Nouvellet P. Estimating the epidemic reproduction number from temporally aggregated incidence data: A statistical modelling approach and software tool. PLOS Comput Biol. 2023; 1–14; 10.1371/journal.pcbi.1011439.

25. Becker NG, Watson LF. A METHOD OF NON-PARAMETRIC BACK-PROJECTION AND ITS APPLICATION TO AIDS DATA. 1991;10: 1527–1542; 10.1002/sim.4780101005

26. Ferguson NM, Cummings DAT, Cauchemez S, Fraser C, Riley S, Meeyai A, et al. Strategies for containing an emerging influenza pandemic in Southeast Asia. Nature. 2005;437: 209–214. doi:10.1038/nature04017

27. Merler S, Ajelli M, Fumanelli L, Parlamento S, Pastore A, Dean NE, et al. Containing Ebola at the Source with Ring Vaccination. PLoS Negl Trop Dis. 2016; 1–11. doi:10.1371/journal.pntd.0005093

28. Warimwe GM, Gesharisha J, Carr BV, Otieno S, Otingah K, Wright D, et al. Chimpanzee adenovirus vaccine provides multispecies protection against rift valley fever. Sci Rep. 2016;6: 1–7. doi:10.1038/srep20617

29. Udahemuka JC, Cassidy H, Schuele L, Uwibambe E, Ngabo MG, Masirika LM, et al. Persistent circulation of Rift Valley fever virus lineage C in Rwanda, 2022–2025. One Heal. 2026;23: 101529. doi:10.1016/j.onehlt.2026.101529

30. Woolhouse, Dye C, Etard JF, Smith T, Charlwood JD, Garnett GP, et al. Heterogeneities in the transmission of infectious agents: implications for the design of control programs. Proc Natl Acad Sci U S A. 1997;94(1):338–342. Available: http://www.ncbi.nlm.nih.gov/pmc/articles/PMC19338/pdf/pq000338.pdf

31. Lloyd-Smith JO, Schreiber SJ, Kopp PE, Getz WM. Superspreading and the effect of individual variation on disease emergence. Nature. 2005;438: 355–359. doi:10.1038/nature04153

32. Lepore L, Odinoh R, Dawa J, Situma S. Understanding the Rift Valley fever exposure risk : A comparative perspective from a multi-country study in East and Central Africa, 2021-24. PLoS Negl Trop Dis. 2026; 1–18. doi:10.1371/journal.pntd.0014082

